# Estimating the number of COVID-19 cases being introduced into UK Higher Education Institutions during Autumn 2020

**DOI:** 10.1101/2020.09.02.20186676

**Authors:** Francisco J. Pérez-Reche, Norval J.C. Strachan

## Abstract

It is estimated that 81% of the 163 UK Higher Educational Institutes (HEIs) have more than a 50% chance of having at least one COVID-19 case arriving on campus when considering all staff and students. Across all HEIs it is estimated that there will be a total of approximately 700 COVID-19 cases (95% CI: 640 – 750) arriving on campus of which 380 are associated from UK students, 230 from international and 90 from staff. This assumes all students will return to campus and that student numbers and where they come from are similar to previous years. According to the current UK government guidance approximately 237,370 students arriving on campus will be required to quarantine because they come from countries outwith designated travel corridors. Assuming quarantining is 100% efficient this will potentially reduce the overall number of cases by approximately 20% to 540 (95% CI: 500 – 590). Universities must plan for COVID-19 cases to arrive on campus and facilitate mitigations to reduce the spread of disease. It is likely that the first two weeks will be crucial to stop spread of introduced cases. Following that, the risk of introduction of new cases onto campus will be from interactions between students, staff and the local community as well as students travelling off campus for personal, educational or recreational reasons.

## Introduction

COVID-19 has resulted in the on-campus closure of HEIs across the UK in March 2020^1^. Since that point universities have been working predominantly as virtual establishments with most staff working from home. Autumn sees the start of the new academic term with the potential return of more than 1.5 million UK and almost half a million international students^2^.

The COVID-19 pandemic continues to run its course across the world with approximately 1000 new cases reported each day in the UK, 25,000 across Europe and 250,000 worldwide^3^ accessed 28/03/20). There have been a number of outbreaks of COVID-19 reported in universities in the USA (The University of North Carolina, Notre Dame in Indiana, Colorado College, Oklahoma State and University of Alabama^4^) where the national infection rate is approximately 10 times higher than the UK^3^. Both UK and Scottish governments are providing advice for HEIs regarding COVID-19 and this is continually evolving^5,6^. However, it is currently unknown to what extent COVID-19 will be brought to campus by staff and students whether from the UK or abroad.

Here we use current estimates of COVID-19 incidence from across the world together with student and staff numbers to determine the most likely number of COVID-19 cases to be introduced to each UK HEI campus at the start of the current term. We determine this by level of occupancy as it is not expected that all students will return to campus. We then determine the likely effect of government quarantine measures for international students and consider campus mitigations to reduce the spread of the virus.

## Materials and Methods

Our analysis considers 163 HEIs in the UK (see Additional File 1). The Open University has been excluded from the analysis since it offers distance learning. The population of any given HEI is split into three subpopulations: Staff, students from UK and international students. For the *i*-th HEI, there are *S_i_* members of staff, *U_i_* students from UK and *I_ij_* international students from each of the *j* = 1,2,… 212 countries of domicile (see the numbers in Additional File 1). These numbers were downloaded from the Higher Education Statistics Agency for the 2018–2019 academic year ^2,7^. Let 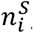, 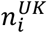 and 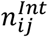 be the number of introduced cases arriving at HEI *i* linked to each of the subpopulations. We assume that these numbers obey binomial distributions: 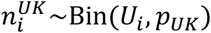, 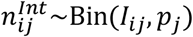 and 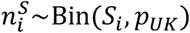. Here, *p_uk_* and *p_j_* are the probabilities that a randomly chosen individual from the UK or country *j* is a COVID-19 case, respectively. Based on the infectivity profile over time for individuals with COVID-19^8^, we regard as potentially infectious cases all those reported in the last 14 days. In particular, we based our estimates on cases reported until 22 August 2020^3^, i.e. the last 14 days used here are those preceding 22 August 2020. We assume the incidence of the virus in this period to be representative of the incidence at the time when students start arriving at HEIs in Autumn 2020. Following this, the ratio between the number of cases in the UK in the last 14 days and the population in the UK gives *p_uk_* = 2.1 × 10^−4^. Similarly, the probability *p_j_* is estimated by the ratio between the number of cases in country *j* in the last 14 days and the population of the country.

Based on the binomial assumption mentioned above, the probabilities that no case arrives at HEI *i* linked to each of the subpopulations are given by

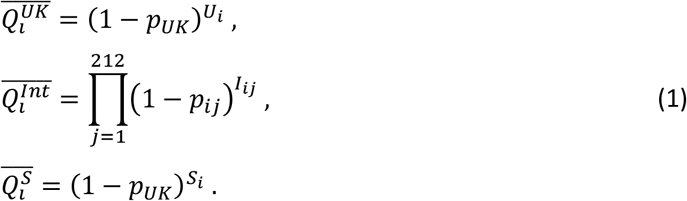

The probabilities that at least one case linked to UK students, international students or staff are then given by 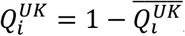, 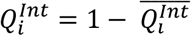 and 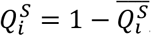, respectively. The probability that there is at least one case at an institution *i* is 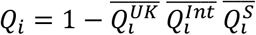

The expected value of the number of introduced cases linked to each of the subpopulations at HEI *i* is given by:

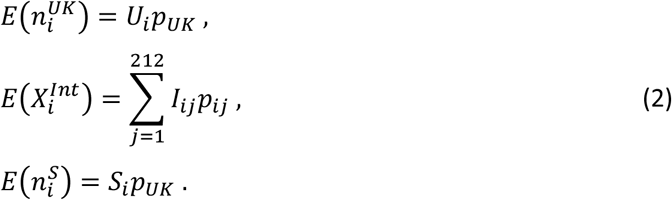

Here, 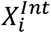 is the total number of cases from international students. The expected value for total number of cases arriving at HEI *i*, 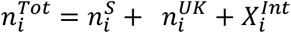 is simply the sum of the expected value given by (Eq. 2).

Scenarios with different occupancies are simulated by reducing the number of UK and international students by a factor *f* ∈ [0,1].

Quarantine of students coming from a country *j* is implemented by effectively setting *I_ij_* = 0 for the country, i.e. we assume that quarantine is 100% efficient. Quarantine is applied to all students with domicile in countries that are not within a UK travel corridor^9^.

## Results and Discussion

Generally, the larger the number of students and staff at an HEI the greater the likelihood of COVID-19 being introduced (Figure 1(a) black symbols). There is more than a 50% chance of at least one COVID-19 case arriving on Campus for institutions with more than ∼3000 students assuming 100% occupancy (i.e. that all students return to campus). This corresponds to 81% of the 163 HEIs. For 65% student occupancy, 78% of institutions would still have a probability larger than 0.5. These are institutions with more than ∼4000 students. Even for a low student occupancy of 30%, the probability of at least one case arriving on campus is greater than 50% for 67% of the HEIs. These are institutions with more than ∼6000 students (green symbols in Figure 1(a)). For most HEIs (127/163) the probability of at least one student introducing COVID-19 to campus is higher for UK students than international students (Fig. 1(b)). This is mainly due to most UK HEIs (97%) having more UK than international students attending.

**Figure 1.**
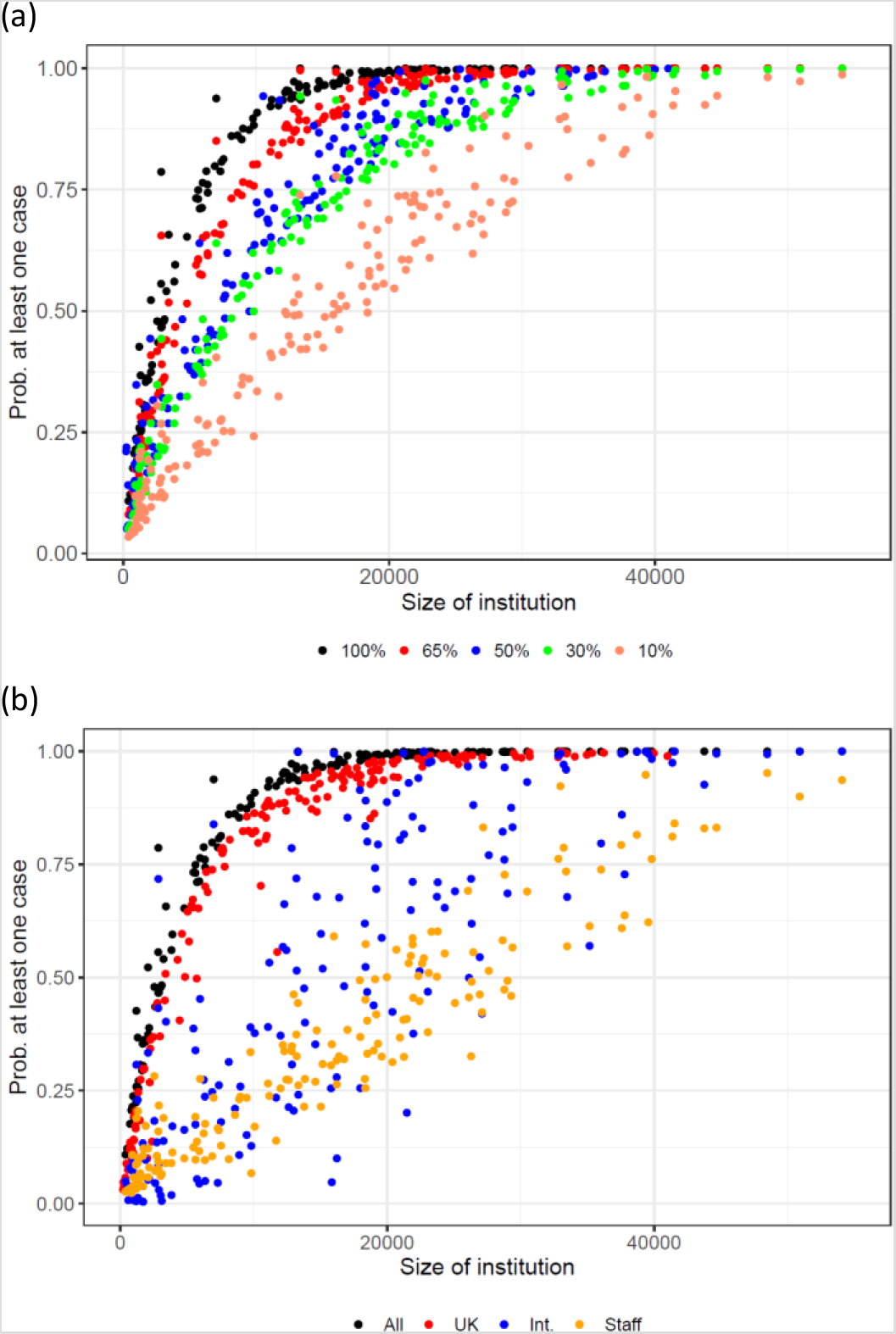
The probability of at least one case of COVID-19 being introduced to campus at the start of term for each institution stratified by (a) level of student occupancy and (b) students (UK or international) and staff assuming 100% occupancy. Note each point represents data from 1 of the 163 UK HEIs. The size of the institutions is the sum of the number of staff and the number of students.

The expected number of introduced cases to campus generally increases with institution size (Figure 2(a)) and numerical values in Additional File 1). In total for all 163 HEIs, an average of approximately 700 COVID-19 cases (95% CI: 640 – 750) would expect to be introduced with 380 and 230 from UK and international students respectively, whilst 90 from staff. The mean number of cases varies from approximately 0.1 cases at The National Film and Television School to approximately 20 at University College London.

**Figure 2.**
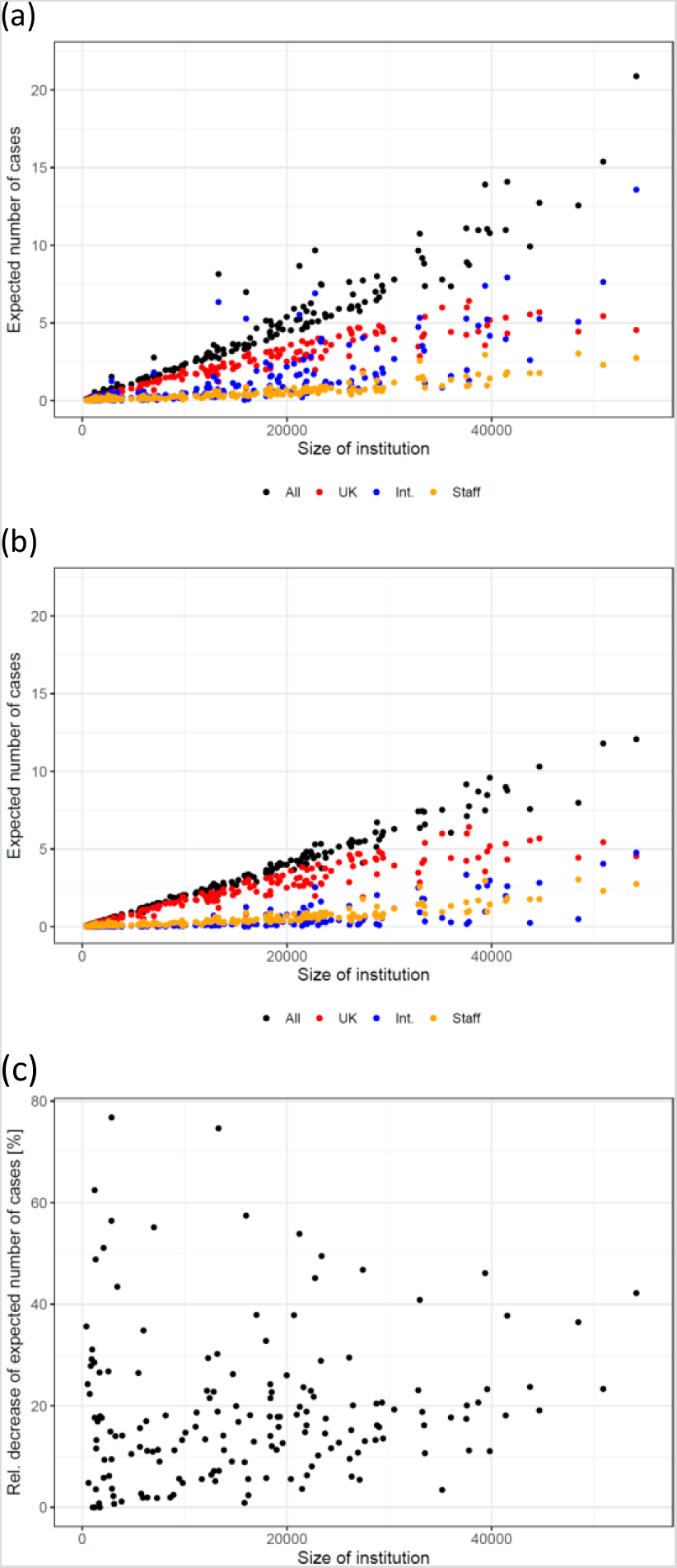
The expected number of cases for each HEI in the UK by size of institution and stratified by (a) staff and student type, (b) the same graph but after the effects of quarantining international students from non-travel corridor countries and (c) the percentage decrease of cases introduced to campus due to quarantine.

Quarantining the 237,370 students living outwith UK travel corridors would lead to a reduction of 154 cases on average (20% overall) with reductions of between 0 to approximately 77% across all HEIs (see panels (b) and (c) in Figure 2 and numerical values in Additional File 1). This assumes that quarantining is 100% efficient. This will likely be a challenge due to students living in multiple occupancy private flats or halls of residence and having to go out to purchase food etc.

The analysis of the average number of cases expected to arrive into an institution gives a first approximation of the risk of outbreaks. Considering the variability of the number of cases in addition to the average, however, is important to properly assess the risk. Figure 3 shows in detail probability distributions for the number of cases arriving on campus, for exemplar small (University of Buckingham), medium (University of Aberdeen) and large (Nottingham Trent University and University College London) institutions in terms of student numbers with and without quarantine of international students. These are estimates for 100% occupancy (results for all HEIs are given in Additional File 3). For large HEIs such as University College London, the predicted number of introduced cases is 21 (95% CI: 12 – 30), i.e. between 12 and 30 cases could arrive on campus (see the black curve in the left panel of Figure 3(d)). Since University College London has a large number of international students, the number of cases might significantly reduce to 12 (95% CI: 6 – 19) if international students that are not within travel corridors quarantine on arrival to the UK (compare the black curves in the left and right panels of Figure 3(d)). The estimated number of cases arriving at the Nottingham Trent University is 9 (95% CI: 4 – 15). Quarantine of international students would only reduce these values to 7 (95% CI: 2 – 13). For the University of Aberdeen, we estimate 5 (95% CI: 1 – 10) cases arriving on campus (left panel of Figure 3(b)). When taking quarantine into account, the predicted number of cases reduces to 3 (95% CI: 0 – 8). For a smaller institution such as the University of Buckingham, the expected number of cases is 1 (95% CI: 1 – 3) without quarantine and 0 (95% CI: 0 – 2) with quarantine.

**Figure 3.**
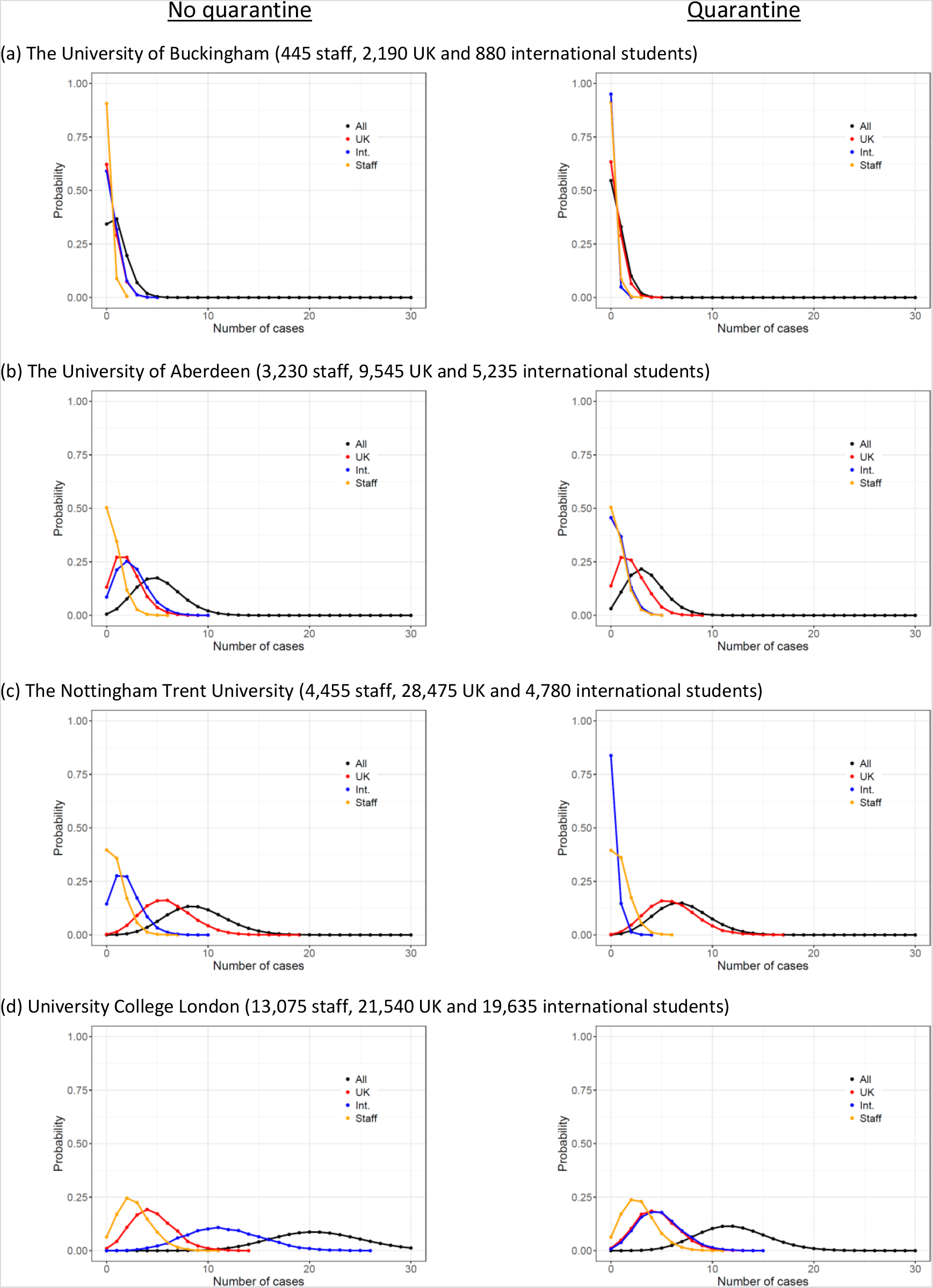
Probability distributions for the number of cases by staff and students type for (a) University of Buckingham (small), (b) University of Aberdeen (medium), (c) The Nottingham Trent University (large) (d) University College London (largest). Panels on the left show results without quarantine. Panels on the right show results with quarantine. All results assume 100% occupancy.

Figure 4 shows the effect of reduced student occupancy on the expected number of cases arriving at the four exemplar institutions selected above. Interestingly, the expected number of cases in relatively large institutions remains high even if occupancy is significantly reduced. For a 65% student occupancy, on average 14.5, 6.1, 3.6 and 0.7 cases would be expected to arrive in University College London, Nottingham Trent University, University of Aberdeen and University of Buckingham. When considering quarantining, these numbers reduce to 8.8, 5.0, 2.5 and 0.4 respectively. Results for all HEIs are given in Additional File 2.

**Figure 4.**
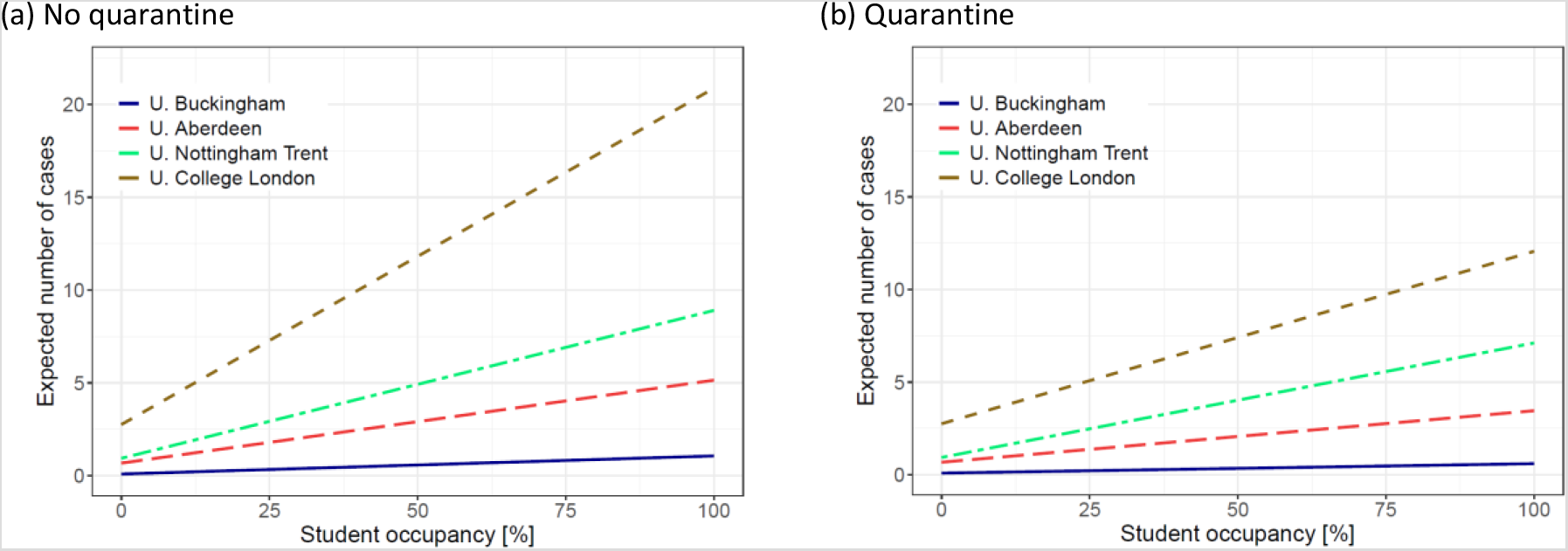
The expected number of cases arriving on campus as a function of student occupancy for selected HEIs assuming (a) no quarantine or (b) quarantine for international students outwith from travel corridors.

The analysis performed here is based on a number of assumptions. First, that all the students attend university in the Autumn. This will be an overestimate because some postgraduate courses have intakes after Christmas and so these students will not be attending in October. Second, it is assumed that the country which the students call home is where they are coming from. Some may have spent the summer in the UK and some will have spent the summer elsewhere. Indeed, some UK students may also have spent the summer abroad. Third, it is assumed that the incidence of COVID-19 is uniform for each country and it is reported accurately. However, it is known that there is a great deal of heterogeneity within a country due to local outbreaks, this can be seen in the UK^10,11^ as well as countries abroad^12–14^. With the available data, it has not been possible to incorporate this heterogeneity into the analysis at a worldwide level. Fourth, the incidence has been estimated based on reported cases. This is likely to underestimate the actual incidence which may include a significant number of unreported cases ^15,16^. Fifth, it is assumed that the data used from 2018/19 will be representative of the student cohort joining HEIs in 2020/21. However, extra funding/places has been made available to HEIs because of the higher grades school pupils have been awarded in the UK due to this being decided on teachers estimates rather than final exams^17^. Also, as a consequence of BREXIT, there may well be fewer new EU students joining UK universities this year than previously due to changes in payment of fees. However, these two changes will only affect new students so should not modify the figures substantially. Hence, because of these assumptions the figures presented must be regarded as estimates and can be refined as more data becomes available.

Many British HEIs plan to offer Blended Learning this academic year which involves a combination of on-campus and on-line teaching^18^.As mentioned above both UK and Scottish governments are providing advice for HEIs^5,6^. Already some universities have released advice for students regards physical distancing, use of face masks, instructions for use of buildings, etc^19^. The independent SAGE group has also suggested that face to face teaching should not occur except for practical training or lab work and have provided a number of recommendations to reduce the risk of COVID-19 transmission on-campus^20^.

The current study which suggests 700 (95% CI: 640 – 750) cases from 212 countries arriving on-campus indicates testing of all students on arrival at campus is impractical since the UK currently has ∼350,000 testing capacity per day^21^. Also, it is estimated that most cases arriving on campus will be from UK students, due to the larger number of UK students attending and that quarantining of international students has the potential to significantly reduce the spread of COVID-19 on campus.

This paper provides results on the potential introduction of COVID-19 onto campus but it does not discuss the potential for spread during term. However, it is likely that the first two weeks returning to campus will be crucial in controlling the spread of this pathogen. “Freshers flu” which comprises a battery of different illnesses (colds, flu etc) has been reported to arise early in term due to proximate mixing of students after the summer break and the emergence of various seasonal respiratory diseases^22,23^. This is also likely to be the case for COVID-19 unless appropriate steps are taken to facilitate its onward transmission. Co-circulation of COVID-19 and other seasonal respiratory diseases which share symptoms with COVID-19 (e.g. colds or flu) poses significant uncertainties on the spread of all these diseases^24^. These uncertainties are not only relevant during the first weeks on campus. Indeed, during term there is the potential for repeated introduction of the virus onto campus due to interactions between students and the local community as well as from staff and their external social networks. The extent to which this will arise will be dependent both on the number of interactions as well as the prevalence in the wider community.

## Conclusion

The results presented here show that most UK HEIs will have at least one COVID-19 case on campus even if only 65% of students return to campus in the Autumn. Quarantining of students outwith travel corridors decreases by 21% the expected number of COVID-19 cases arriving on campus but the chances that there will be at least one case are not very much affected by quarantining. Hence, it will be prudent for HEIs to plan for COVID-19 cases to arrive on campus and facilitate mitigations to reduce the spread of disease. It is likely that the first two weeks of term will be crucial to stop spread of introduced cases. Following that, the risk of introduction of new cases onto campus will be from interactions between students, staff and the local community as well as students travelling off campus for personal, educational or recreational reasons.

## Data Availability

All data sources have been referred in the manuscript

https://ourworldindata.org/covid-cases

https://www.hesa.ac.uk/data-and-analysis/students/where-from

https://www.hesa.ac.uk/data-and-analysis/staff/table-1

## Author contributions statement

FPR and NS planned the project and designed the research. FPR retrieved the data and did the statistical analysis. NS and FPR wrote the manuscript.

## Additional files

Additional file 1. Population of HEIs and expected number of COVID-19 cases introduced to each institution this autumn. The expected number of cases from international students is given both with and without quarantine (see last two columns). Results provided assume 100% occupancy.

Additional file 2. Population of HEIs and expected number of COVID-19 cases introduced to each institution this autumn. The expected number of cases from international students is given both with and without quarantine (see last two columns). Results provided assume 65% occupancy.

Additional file 3: Summary statistics (2.5% percentile, median, 97.5% percentile) of the probability distribution of the number of cases being introduced into each HEI with and without quarantine. Results assume 100% occupancy.

